# THE ROLE OF KNOWLEDGE AND ATTITUDE ON HIV AND AIDS PREVENTION PRACTICES AMONG SECONDARY SCHOOL STUDENTS: A CROSS-SECTIONAL STUDY OF GWASSI SOUTH SUB-COUNTY, HOMA BAY COUNTY, KENYA

**DOI:** 10.1101/2023.01.10.23284403

**Authors:** Kennedy Odhiambo Akello, Japheths Ogendi, Collins Otieno Asweto

## Abstract

**Background:** Homa Bay County HIV prevalence is the highest and 4.5 times the national prevalence. Young people aged 15-24 years account for 22% of this prevalence in the County. Majority of these young people are high school students, yet their knowledge, attitudes, practices and risk perception towards HIV prevention have not been unascertained. This could be one of the obstacles to HIV prevention in the county.

**Objective:** The study aimed at assessing HIV-related Knowledge, Attitudes and Practices (KAPs) of high school students in Homa Bay County.

**Methodology:** Survey was conducted among 260 systematically sampled students from 11 randomly selected schools in Gwassi South ward, Suba South sub-County, Homa Bay County. A head teacher or health master from each of the 11 schools was purposively sampled and took part in key informants’ interview about available strategies for HIV prevention. Data was analyzed using SPSS version 18.0. Proportion was used to determine knowledge, attitude, and risk perception among the students. Logistic regression was used determine association between knowledge, attitude towards HIV & AIDS and sexual practices.

**Results:** Majority (64%) of students had moderate to high knowledge on HIV & AIDS and exhibited positive attitude towards HIV & AIDS (61.1%). However, 15% to 25% of students reported to have engaged in a risky sexual practice. Higher to moderate knowledge had increased likelihood of positive attitude towards HIV and AIDS. Similarly, those with higher to moderate knowledge demonstrated lower risk sex practices. Moreover, students with negative attitude towards HIV & AIDS were 4 times more likely to have high risk sex practices. There were HIV & AIDS intervention strategies in secondary schools within Gwassi South Ward.

**Conclusion:** Knowledge and attitude of the students play a big role in their HIV risk perception as well as sex practices. Understanding knowledge, attitude, practice and intervention strategies of HIV and AIDS is significant in policy formulation by Ministries of Education, and leads to improved health-related practice by the Ministry of Health and researchers focusing on HIV and AIDS in high burden area.

## Background Information

Globally, approximately 40.1 million have died due to Human Immune-deficiency Virus (HIV) and acquired Immune Deficiency Syndrome (AIDS) while 38.4 million people were reported to be living with the disease, by 2021 (WHO, 2022). The HIV epidemic has disproportionately affected people in Sub-Saharan Africa, particularly children, given that 88% of HIV-positive children and adolescents were living in sub-Saharan Africa including Kenya as of 2021 (UNICEF, 2022a). Approximately 1.4 million people were reported to be living with HIV infection in Kenya by the year 2020 (UNAIDS, 2022). HIV and AIDs also contributes to a sizeable health and economic burden in Kenya. For example, HIV and AIDS accounts for an estimated 29% of annual mortality amongst adults; 20% of maternal mortality, and; 15% of deaths of children under the age of five years, and also affects the economy adversely (Ministry of Health, 2016).

The Global Youth Wellbeing Index, which includes the component of health, reveals that 85% of youth (age 10 to 24) in the 30 countries sampled, report low levels of overall well-being (Center for Strategic & International Studies, 2014). HIV and AIDS is one of the health problems that disproportionately affects the youth. For example, HIV mortality is reported to be increasing in the adolescent age category, even though in other sub-populations, HIV mortality has shown downward trend (Berker, Johnson, Wallace, & Hosek, 2015). A growing proportion of HIV-positive individuals worldwide are adolescents and young people. About 0.41 million young people between the ages of 10 and 24 were newly infected with HIV in 2021, of whom 0.16 million were teenagers between the ages of 10 and 19 (UNICEF, 2022b). To make matters worse, most recent data show that only 25% of adolescent girls and 17% of adolescent boys aged 15-19 in Eastern and Southern Africa - the region most affected by HIV - have been tested for HIV in the previous 12 months and have received the results of the most recent test. Given the current trend, approximately 0.18 million new HIV infections will be reported annually among adolescents each year in 2030 (UNICEF, 2022b). The increase in HIV deaths in the adolescent age category has occurred mainly in the African region, resulting in AIDS being the leading cause of deaths among adolescents in Africa, and the second leading cause for deaths amongst adolescents worldwide (World Health Organization, 2014).

In Kenya, Homa Bay County is the leading County nationally in HIV prevalence. The HIV prevalence of 19.6% in the county in 2018 was 4 times higher than the national HIV average prevalence of 4.9% (Achia et al., 2022). The County is also the second leading nationally, in terms of the number of people living with AIDS and contributed 10.4% of the total number of people living with HIV in Kenya by the end of 2015, 22% of which were young people aged 15-24 years and 6% being children under the age of 15 years (Mandiwa, Namondwe, & Munthali, 2021).

The Vision indicated in the First Homa Bay County Multi-Sectoral Aids Strategic Plan is “A County with the highest possible standards of health, in a gender sensitive environment, free from HIV, stigma and discrimination” and with a Mission “to design and provide integrated, devolved, rights-based, evidence-informed and cost-effective HIV interventions and services that ensure timely HIV interventions”. However, high drop-out rate out of learning institutions by young girls and boys to engage in manual work and early marriage have been identified as some of the gaps to be addressed in HIV programming in the County (County Government of Homa Bay, 2013b). There has also been concern about the knowledge, attitude, practice and risk perception amongst the youth in Suba sub County by the sub-County health authorities (Otieno, Karanja, & Kagira, 2018).

Previous studies amongst the youth, pointed out that condom use was low among the youth and adolescents and that the youth still held myths and misconceptions about HIV and AIDS (Wairimu, 2014). It was also shown that young people continue to accept favors such as gifts in exchange of sex due to their poor risk perception of contracting the disease (Ithibu, 2015). These studies, however, were conducted in urban settings of the country.

Gwassi sub-County being greatly inaccessible due to poor geographical terrains and poor roads remain cut off from information and hence adolescents and youth bear the burden of information access. Previous studies conducted in Gwassi reveal that adolescents lack accurate information or knowledge on HIV and AIDS. Their sexual behaviors and social conditions also expose them to the risk of acquiring HIV and AIDS (Ooyi, 2015).

The main objective of this study was to assess HIV and AIDS knowledge, risk perception, and attitudes toward safer sex practices among students enrolled in secondary schools in Gwassi South Ward in Suba South sub-County of Homa Bay County. Suba South sub-County is amongst the top three leading sub-Counties in terms of HIV prevalence (NASCOP, 2015). The study was based in Gwassi South Ward which is one of the four (4) administrative wards in the sub-County.

Understanding the knowledge, attitude and sexual practice among the secondary school students is significant because the various developmental, psychological, social, and structural transitions that converge in this period of the lifespan render the sub-population more vulnerable to HIV (Berker et al., 2015). Furthermore, young people are the world’s greatest resource (Berker et al., 2015). Reaching adolescents is very crucial for public health strategies aimed at HIV prevention and decreasing HIV-related deaths.

## METHODOLOGY

### Study Site

The study was conducted in Gwassi South ward of Suba South sub-County in Homa Bay County. Homa Bay County covers an area of 4,267.1 Km2 inclusive of the water surface and lies between latitude 0°15’ South and 0°52’ South, and between longitudes 34°East and 35°East (County Government of Homa Bay, 2013a). The county is located in South Western Kenya along Lake Victoria and boarders Kisumu and Siaya counties to the North, Kisii and Nyamira counties to the East, Migori County to the South and Lake Victoria and the Republic of Uganda to the West (County Government of Homa Bay, 2013a). Gwassi South is one of the five administrative wards in Suba South sub-County, namely (Republic of Kenya, 2013): Gwassi South, Gwassi North, Kaksingri West, Ruma and Kaksingri East. Gwassi South Ward borders Migori County to the South, Lake Victoria to the West and Gwassi North to the North.

### Study Design

This was a cross-sectional study in which structured questionnaires were used to collect data on knowledge, attitude and practice towards HIV and AIDS prevention among secondary students enrolled in schools in Gwassi South Ward. Qualitative data was collected among head teachers/health masters using Key Informant Interview guide.

### Study Population

The study population is comprised of students enrolled in secondary schools in Gwassi South Ward, during the study period and principals and teachers charged with the responsibility of students’ health in the sampled schools.

### Inclusion

Respondents must be a secondary school student in Gwassi South Ward, Homa Bay County, and willing to participate into the study and be able to provide written informed consent/assent.

### Sample Size Determination

The sample design of this study was calculated based on Cochran (1963): **n**=z^2^pq/d^2^. Where; n = desired sample size, z = standard normal deviate at the required confidence level, d = the margin of error allowed, which is 5% in this survey (0.05), or degree of accuracy desired is 95%, p = proportion of the target population or estimated characteristics being measured (0.23), q = estimated characteristics not being measured (1-0.5 = 0.5)’ Substituting, = (1.96) ^2^ (0.23) (0.77)/0.05^2^ = 272. Because the population was less than 10,000, the following correction formula was applied: nf =n/[1+(n/N)], giving = 272/ (1+(272/6400) = 260. In order to cater for non-response, 10% of 260 (which is 26) were added to 260 making the final sample size proposed to be 286.

### Sampling Techniques

Sampling of participants in secondary schools in Gwassi South ward was conducted according to probability proportional to size (PPS) (Cheung, 2014). This is a technique, in which the probability of selection for each sampling unit in the population is proportional to an auxiliary variable (Thomas et al., 2002). This sampling technique is used in <u>survey research</u> when the sampling units vary in size or in other important aspects that the researchers want to take into account in the sample design (Makela, Si, & Gelman, 2018), as indicated in table 3.1 below. In each school, systematic random method was used to get the desired number of participants.

**Table 1:** Probability Sampling based on Population of secondary students in Gwassi South ward; October 2019.

| No | School | Number of students per school | Proportion of total (%) | No. Sampled (n) |
| --- | --- | --- | --- | --- |
| 1 | God Oloo | 601 | 9 | 26 |
| 2 | God Bura | 650 | 10 | 29 |
| 3 | Homa Secondary | 550 | 9 | 26 |
| 4 | Kagoro Girls | 652 | 10 | 29 |
| 5 | Kiabuya | 550 | 9 | 26 |
| 6 | Kigoto Secondary | 599 | 9 | 26 |
| 7 | Magunga | 647 | 10 | 29 |
| 8 | Nyatembi | 498 | 8 | 23 |
| 9 | Olando | 551 | 9 | 26 |
| 10 | Seka | 502 | 8 | 23 |
| 11 | Wiga Secondary | 600 | 9 | 26 |
| **TOTAL** |  | **6400** | **100** | **286** |

Gwassi South Ward was randomly picked for this study. Five similar papers were marked with the names of all the five Wards in Suba Sub County. They were put in a small basket and shaken to ensure that they were thoroughly mixed up. One person who was not involved in writing the names was told to pick one paper. That picked ward would be the ward in which the study would be conducted and it’s Gwassi South Ward that was picked.

#### Data Collection Instruments

### Questionnaires

The questionnaire used in the study was developed by the researcher in consultation with experts in HIV and AIDS field. Previous related HIV work from Family Health International’s questionnaire on HIV and AIDS prevention in developing countries (Project-USAID, 2003), Kenya’s National Aids and STIs Control Programme (NASCOP, 2015) and a local study on HIV knowledge, attitude and practice (Salmen et al., 2015) were consulted and applied in developing the questionnaire. The final questionnaire used in this study had questions relating to HIV knowledge, attitudes toward PLHIV and sexual practices, in addition to socio-demographic information. The questionnaire was divided into four sections, A, B, C and D. Section A focused on the socio-demographic characteristics of the participants, capturing age, sex, and religion.

Section B had 18 knowledge-based items. Section C comprised 17 questions on attitudes towards PLHIV. Finally, Section D comprised 7 questions that focused on behavior/practices related to HIV and AIDS. In order to assess the clarity, feasibility, and appropriateness of the questionnaire for the students, the questionnaire was piloted with 11 students in within the study area. This proportion had previously been used in a study focusing in HIV knowledge, attitude and practice in low and middle income country (Thanavanh, Harun-Or-Rashid, Kasuya, & Sakamoto, 2013). The questionnaire was developed in English and there was no need for translation since the students registered in Kenya are taught in English and use English as a medium of communication.

### Interview Schedule for Head Teachers

In order to solicit detailed information, key informant interviews were conducted among head teachers of public secondary schools in Gwassi South Ward. A semi-structured guide with open-ended questions was used in the data collection process. This gave the researcher an opportunity to seek more clarification on a research question regarding which strategies would best work in preventing HIV and AIDS among this population. A key informant interview schedule is an important tool for gathering data as the interview situation allows much greater depth than other methods of data collection (Cohen et al., 2017). It attempted to provide a true picture of opinions and feelings from key people who were more informed about the behaviour characteristics of the study population.

#### Measurement of Variables

The information gathered from participants included demographic characteristics; knowledge on; attitude towards; and sexual practices (risk perception) towards HIV and AIDS among students; as well as availability and implementation of HIV prevention intervention strategies in secondary schools in Gwassi South ward in Suba South sub-County.

### Independent variables

The independent variables in this study include knowledge, attitude (cognition, affect, and behavior) and risk perception towards HIV and AIDS among the students in Gwassi South Ward in Suba South sub-County. To determine the respondents’ level of HIV-related knowledge, they were read for 18 HIV-related statements with options as ‘‘true’’, ‘‘false’’ or ‘‘do not know’’ to every knowledge related question. A score of **“1”** was assigned for a correct answer and **“0”** for a wrong answer or a “do not know” response. The scores were then summed up to generate an overall score of each participant. The level of knowledge was then categorized into “*low*” for respondents scoring ≤ 50%, “*moderate*” for those scoring between 51% and 74% and “*high*” for scores ≥ 75%.

For attitude, a 17-statement questionnaire was administered and participants to answer “Yes” or “No” based on what they feel. Those scoring less than mean scores were classified as having ‘‘negative’’ and those scoring equal to or more than mean scores were classified as having ‘‘positive’’ attitudes.

### Dependent variable

The dependent variable in this study is safe sex practices as measured by the prevalence of preventive behaviors exercised by the respondents (students in Gwassi South ward in Suba South sub-County). Such preventive behaviors include use of condoms, abstinence, being faithful to one partner and uptake of VCT services.

### Data Collection Procedures

Before data collection began, students were briefed about the purpose of the study and technical terminologies used in the questionnaire. Guidance was given on how to fill out the questionnaire form. Written informed assent was obtained for all the students surveyed prior to administering the questionnaires. Those who agreed to participate were met in the classroom by the researchers who then distributed the questionnaires to be self-administered. This was done during the 30 minutes break of their class. The students were assured that their responses would be treated confidentially. Respondents were also informed that their participation was entirely voluntary and that they were free not to respond to any question that they were uncomfortable with. The head teacher at each engaged school was approached in order to seek permission prior to conducting the surveys.

### Piloting of Study Tools

In order to assess for clarity, feasibility, and appropriateness of the questionnaire for the students, the questionnaire was piloted with 11 students in neighboring schools in Kaksingri Central ward. This ward was chosen due to its demographic and geographic similarity with Gwassi South ward. This proportion has previously been used in a study focusing in HIV knowledge, attitude and practice in low and middle income country (Thanavanh et al., 2013). The tools were found to be reliable and valid except for a few adjustments on logical flow which were made.

### Data Handling and Management

Completed data forms were thoroughly reviewed to ensure completeness, accuracy and logical flow. Fully reviewed data was entered into Excel spreadsheet and exported to SPSS. Coding was done at the entry stage to ensure all categorical variables are stored as coded data and labels are attached to each code. Both data form in source documents and Excel spreadsheet were password protected and stored confidentially-only the lead researcher and assistant were able to access.

### Validity of research instruments

To test content validity, the researcher sent out test papers to experts to give clarity whether the tests actually measured what it was intended to. Expert opinions were of great help to establish content validity. As such, the researcher sought assistance from supervisors and other experts from the university. All relevant expert opinions were incorporated into the questionnaire during review and so the instrument finally had valid content.

### Data Analysis

Data were entered into an Excel spreadsheet and exported to Statistical Package for the Social Science (SPSS) for Windows, version 18.0 software (SPSS Inc., Illinois, USA) for analysis. Descriptive statistics were to summarize and demonstrate demographic characteristics, knowledge, attitude and practice of students about HIV and AIDS. Categorical data were presented in numbers and percentages. For normally distributed continuous data, the mean (standard deviation, SD) was used while median (interquartile range, IQR) was used for non-normal continuous data. A logistic regression model was used to determine association between level of knowledge on and attitudes towards HIV and AIDS and sex practices. Odds ratios (OR) were calculated at 95% confidence intervals (CIs) and a p-value of ≤ 0.05 was considered significant.

To evaluate knowledge and attitude of the students, they were asked to reply with ‘‘yes’’, ‘‘no’’ or ‘‘don’t know’’ to every knowledge- and attitude-related question. For practices, ‘‘yes’’ or ‘‘no’ ‘options were used. A score of 1 was assigned for a correct answer and 0 for a wrong answer for knowledge- and practice-related questions; for attitude section, it was 1 for every positive answer and 0 for negative ones. The scores were then summed up to generate an overall score for each participant. Levels of KAPs were then re-categorized depending on their total, mean and median score.

Accordingly, level of knowledge was categorized into ‘‘low’’ for respondents scored ≤50%, ‘‘moderate’’ for those scored between 51 and 74%, and ‘‘high’’ for those who scored ≥75%. This categorization was adopted from a study that previously described correlates of misperceptions in HIV knowledge and attitude towards people living with HIV and AIDS among in-school and out of school adolescents in Ghana (Sallar, 2009; Tarkang, Lutala, & Dzah, 2019).

The scores of attitudes and practices were categorized into two segments based on their mean and median score: those scoring less than mean scores for attitude were classified as ‘‘negative’’ and those scoring equal, and more than mean scores were classified as ‘‘positive’’ attitudes. As data for practice was not normally distributed, we used median as the cut-off. Accordingly, those scoring less than median scores for practice were classified as ‘‘risky’’ practices, and those scoring equal, and more than median scores were classified as ‘‘safe’’

### Ethical Considerations

Authorization for study was sought and obtained from Maseno University School of Graduate Studies (SGS). Ethical Clearance was sought and obtained from Maseno University Ethical Review Committee (MUERC) (MSU/DRPI/MUERC/00689/19). Permissions to conduct the study in the County was sought and obtained from County and sub-County Directors of Education and Health. Informed consent was sought and obtained from respective heads of schools on behalf of students. Recognizing that majority of the students are emancipated minors, assent was sought for students aged 13 to 17 years from the head teachers. Students aged above 18 years directly provided informed consent. Respondents were assured that the information given would only be used for the research purpose and treated with utmost confidentiality. Moreover, the respondents were asked not to indicate their names on the questionnaires to ensure anonymity of their responses. Further, confidentiality of information was ensured through secured data storage-data collected was stored in soft copy and password protected and would be backed up in cloud for at least 5 years after study completion. No participant was forced to participate in the study, neither were they unduly influenced.

## RESULTS

### Socio-demographic Characteristics

A total of 260 students took part in the study. Of these number, 115 (44.2%) were male and 145 (55.8%) were female. The ages ranged from 13 to 20 years, with mean age of 17.2 years (SD±1.6). Nearly all participants, 254 (97.7%), were Christians (Table 2).

**Table 2:**
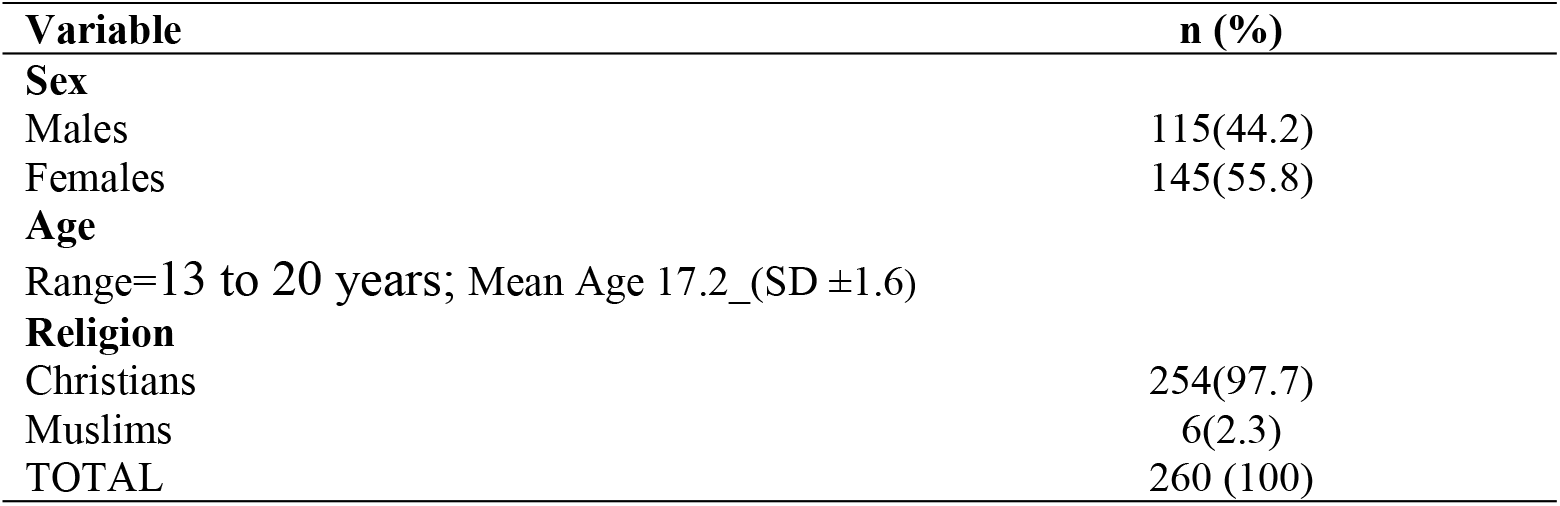
Socio-demographic characteristics of study participants.

| Variable | n (%) |
| --- | --- |
| **Sex** |  |
| Males | 115(44.2) |
| Females | 145(55.8) |
| **Age** |  |
| Range=13 to 20 years; Mean Age 17.2_(SD ±1.6) |  |
| **Religion** |  |
| Christians | 254(97.7) |
| Muslims | 6(2.3) |
| TOTAL | 260 (100) |

### Knowledge on HIV and AIDS

To evaluate the respondents’ HIV-related knowledge, participants were read for 18 HIV-knowledge related statements with options as ‘‘true’’, ‘‘false’’ or ‘‘do not know’’ to every knowledge related question. We assigned a score of **“1”** for a correct answer and **“0”** for a wrong and “do not know” responses. The scores were then summed up to generate an overall score of each participant. The mean score was 11.8 (SD±2.7), with a range score of 3-17. The level of knowledge was then categorized into “low” for respondents scoring ≤ 50% (score ≤9), “moderate” for those scoring between 51% and 74% (9 < score ≤11) and “high” for those who scored ≥ 75% (score>11).

### Knowledge on HIV/AIDS transmission

Table 3 presents the findings on knowledge on HIV transmission and prevention from the participants of the study. Overall, the knowledge about route of transmission of HIV was high for some factors and relatively low for other factors. The findings of this study indicate that, 93.1% rightfully knew that it was false that a person can get HIV by sharing a glass of water with someone who has HIV. The majority of students (93.1%) also rightly indicated that the statement that “a person can get HIV by bathing in the same place as a person who has HIV” was untrue. However, there was confusion about some routes of transmission. For example, only 40.8% knew that a woman can get HIV if she has anal sex with a man. There was also confusion regarding the question that asked whether pulling out the penis before a man gets his climax keeps a woman from getting HIV during sex; only 48.1% gave the correct response. Regarding the question on whether a person can get HIV from oral sex, about a half (51.2%) had the correct answer.

**Table 3:** Knowledge on HIV route of transmission, prevention and control; secondary school students.

| No | Question | n (%) |
| --- | --- | --- |
| ***Knowledge about route of transmission*** |  |  |
| 1 | Coughing and sneezing DO NOT spread HIV (True) | 190 (73.1) |
| 2 | A person can get HIV by sharing a glass of water with someone who has HIV (**False**) | 242 (93.1) |
| 3 | Pulling out the penis before a man climax / cums keeps a women from getting HIV during sex (**False**) | 125 (48.1) |
| 4 | A woman can get HIV if she has anal sex with a man (True) | 106 (40.8) |
| 5 | Showing or washing one's genitals/private parts, after sex keeps a person from getting HIV (False) | 199 (76.5) |
| 6 | All pregnant women infected with HIV quickly show serious signs of being infected (**False**) | 179 (68.9) |
| 7 | People who have been infected with HIV quickly show signs of being infected (False) | 186 (71.5) |
| 8 | People are likely to get HIV by deep kissing, putting their tongue in their partner's mouth, if their partner has HIV (**True**) | 212 (81.5) |
| 9 | A woman cannot get HIV if she has sex during her periods (False) | 197 (75.8) |
| 10 | Having sex with more than one partner can increase a person's chance of being infected with HIV (**True**) | 171 (65.8) |
| 11 | A person can get HIV by bathing in the same place as a person who has HIV (**False**) | 242 (93.1) |
| 12 | A person can get HIV from oral sex (**True**) | 133 (51.2) |
| ***Knowledge about prevention and control*** |  |  |
| 13 | There is a vaccine that stops adults from getting HIV (**False**) | 170 (65.4) |
| 14 | There is a female condom that can decrease a woman's chance of getting HIV (**True**) | 180 (69.2) |
| 15 | A natural skin condom works better against HIV than does a latex condom (**False**) | 95 (36.5) |
| 16 | A person will NOT "feel" HIV if she or he is taking antibiotics (**False**) | 131 (50.4) |
| 17 | Taking a test for HIV one week after having sex will tell a person if she or he has HIV (**False**) | 150 (57.7) |
| 18 | Using Vaseline or baby oil with condoms lowers the chance of getting HIV (**False**) | 165 (63.5) |

### Knowledge on HIV/AIDS prevention

Table 3 also summarizes the knowledge of the students about prevention of HIV. A moderate level of knowledge was reported by students when they were asked questions on: whether there is a female condom that can decrease a woman’s chance of getting HIV (69.2%); whether there is a vaccine that stops adults from getting (65.4%) and; whether using Vaseline or baby oil with condoms lowers the chance of getting HIV (63.5%).

However, in response to the question whether “a natural skin condom works better against HIV than a latex condom”, only 36.5% gave the correct response. In, response to a statement “A person will NOT “feel” HIV if she or he is taking antibiotics”, only 50.4% rightly stated that this was false statement.

### Overall Knowledge on HIV and AIDS on Transmission routes and Prevention

Table 4 presents the level of knowledge on HIV/AIDS. Overall, respondents had a mean (11.8 (SD± 2.7), with a range score of (3–17) from score of 18 knowledge-related questions. Accordingly, 45% were classified as having a high level of knowledge (score of ≥75%; 19% as having medium level of knowledge (scores of 51% to 74%), and 36% as having low level of knowledge (scores of ≤50%) regarding HIV/AIDS.

**Table 4:** Knowledge on HIV/AIDS among secondary students in Gwassi South.

| Knowledge Category | Category Score | n (%) |
| --- | --- | --- |
| Low | (Less than) $\leq 50\%$ | 94 (36%) |
| Medium | Between 51% - 74% | 49 (19%) |
| High | (More than) $\geq 75\%$ | 117 (45%) |

### Attitude

The scores of attitudes were normally distributed with a mean score of 10.2 (SD±3.1); minimum score was 2 while maximum score was 17. The scores were therefore categorized into two segments based on the mean score: those scoring less than mean score (<10.2) for attitude were classified as having ‘‘negative’’ and those scoring equal to or more than mean scores (≥10.2) were classified as having ‘‘positive’’ attitudes. The table below gives frequency of the HIV & AIDS attitude score (Table 5).

**Table 5:** HIV and AIDS Attitude (n=260)

| Qs. | Statement on HIV & AIDS Attitude | Positive Attitude<br>n (%) | Negative Attitude<br>n (%) |
| --- | --- | --- | --- |
| 1 | Most people think that getting HIV is a punishment for bad behavior | 187 (71.9) | 73 (28.1) |
| 2 | Most people would not sit next to someone with HIV in public or private transport | 223 (85.8) | 37 (14.2) |
| 3 | Most people think that having HIV is just a matter of bad luck | 170 (65.4) | 90 (34.6) |
| 4 | Most people think less of someone because they have HIV | 138 (53.1) | 122 (46.9) |
| 5 | Most people would not like someone with HIV to be living next door | 202 (77.7) | 58 (22.3) |
| 6 | Most people would reject the friendship of someone with HIV | 106 (40.7) | 154 (59.3) |
| 7 | Most people feel that it is safe for a person with HIV to look after somebody else's children | 70 (26.9) | 190 (73.1) |
| 8 | Most people think that people with HIV can teach us a lot about life | 153 (58.9) | 107 (41.1) |
| 9 | Most people would not date a person if they know that he / she has HIV | 73 (28.1) | 187 (71.9) |
| 10 | Most people are afraid to be around people with HIV | 159 (61.2) | 101 (38.8) |
| 11 | Most people feel that if you have HIV it is your fault | 131 (50.4) | 129 (49.6) |
| 12 | Most people feel that people with HIV deserve as much as anyone else | 133 (51.1) | 127 (48.9) |
| 13 | Most employers would not hire someone with HIV to work for them | 175 (67.3) | 85 (32.7) |
| 14 | Most people would not drink from a tap if a person with HIV had just drunk from it | 181 (69.6) | 79 (30.4) |
| 15 | Most people believe that if you have HIV you must have done some wrong to deserve it | 171 (65.8) | 89 (34.2) |
| 16 | Most people believe that someone with HIV should be ashamed of themselves | 208 (80.0) | 52 (20.0) |
| 17 | Most people feel uncomfortable around people with HIV | 171 (65.7) | 89 (34.3) |

In summary, majority (61.1%) of the respondents exhibited positive attitude towards HIV and AIDS as shown below (Table 6).

**Table 6:** HIV and AIDS Attitude Categorization (N=260)

| HIV attitude Categorization | Frequency | Percentage |
| --- | --- | --- |
| Negative Attitude (scores <10) | 101 | 38.9% |
| Positive Attitude (Scores $\geq 10$) | 159 | 61.1% |
| TOTAL | 260 | 100% |

### Association of knowledge about HIV and AIDS with attitude

In order to determine the relationship between level of knowledge on HIV and AIDS and safer sex practices among secondary school students, a binary logistic regression was performed. The ‘attitude’ status, being the dependent variable, was coded as a binary outcome, with “positive attitude” coded as “0” and “negative attitude” coded as “1”. The knowledge about HIV in categories of “low”, “moderate” and “high” was our independent variable (Table 7).

**Table 7:**
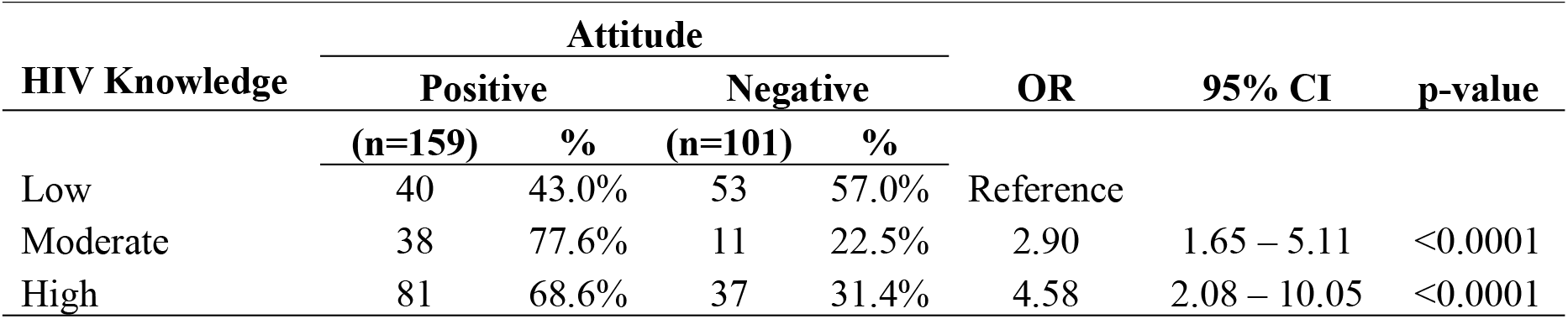
Association of knowledge about HIV and AIDS and Attitude.

As shown in the table above, it was found that level of knowledge was associated with attitude towards HIV and AIDS (p<0.0001).

### Sex Practices

A total of 39 (15%) of the respondents indicated that they had a history of having sexual intercourse with more than one person within six months prior to the survey (Table 8). Slightly less than a quarter, 55(21.2%) indicated that they had engaged in sex without condom with someone who was not their regular sex partner. A quarter (25%) of the students revealed that they had engaged in sex without condom, with someone whose HIV status they did not know (Table 8). Regarding sex after intoxication with alcohol, 8(3.1%), admitted that they had practiced sex directly after consuming alcohol.

**Table 8:** Sex Practices (sex risk behaviour) of the respondents (n=260)

| **Sexual risk behaviour** | **n (%)** |
| --- | --- |
| Had sex with more than one person in the last 6 months | 39 (15.0) |
| Ever had sex without a condom with someone not their regular sex partner | 55 (21.2) |
| Ever had sex directly after consuming alcohol | 8 (3.1) |
| Ever had sex in exchange for money, goods or favour. | 35 (13.5) |
| Ever had sex without a condom with someone whose HIV status they don't know | 65 (25.0) |
| Ever had sex without a condom with someone they knew was HIV positive | 23 (8.8) |
| Ever had sex with someone who is younger or older than them by 15 years | 25 (9.6) |
| Thinking about choices over the last 6 months, rated a high risk of being infected with HIV | 36 (13.9) |

As data for practice was not normally distributed, we used median as the cut-off. Accordingly, those scoring less than median scores for practice were classified as having ‘‘safe’’ sexual practices, and those scoring equal and more than median scores were classified as having ‘‘risky’’ sexual practices. Figure 2 (below) further illustrates the HIV risk score of the respondents as derived from their sexual practices.

**Figure 1:**
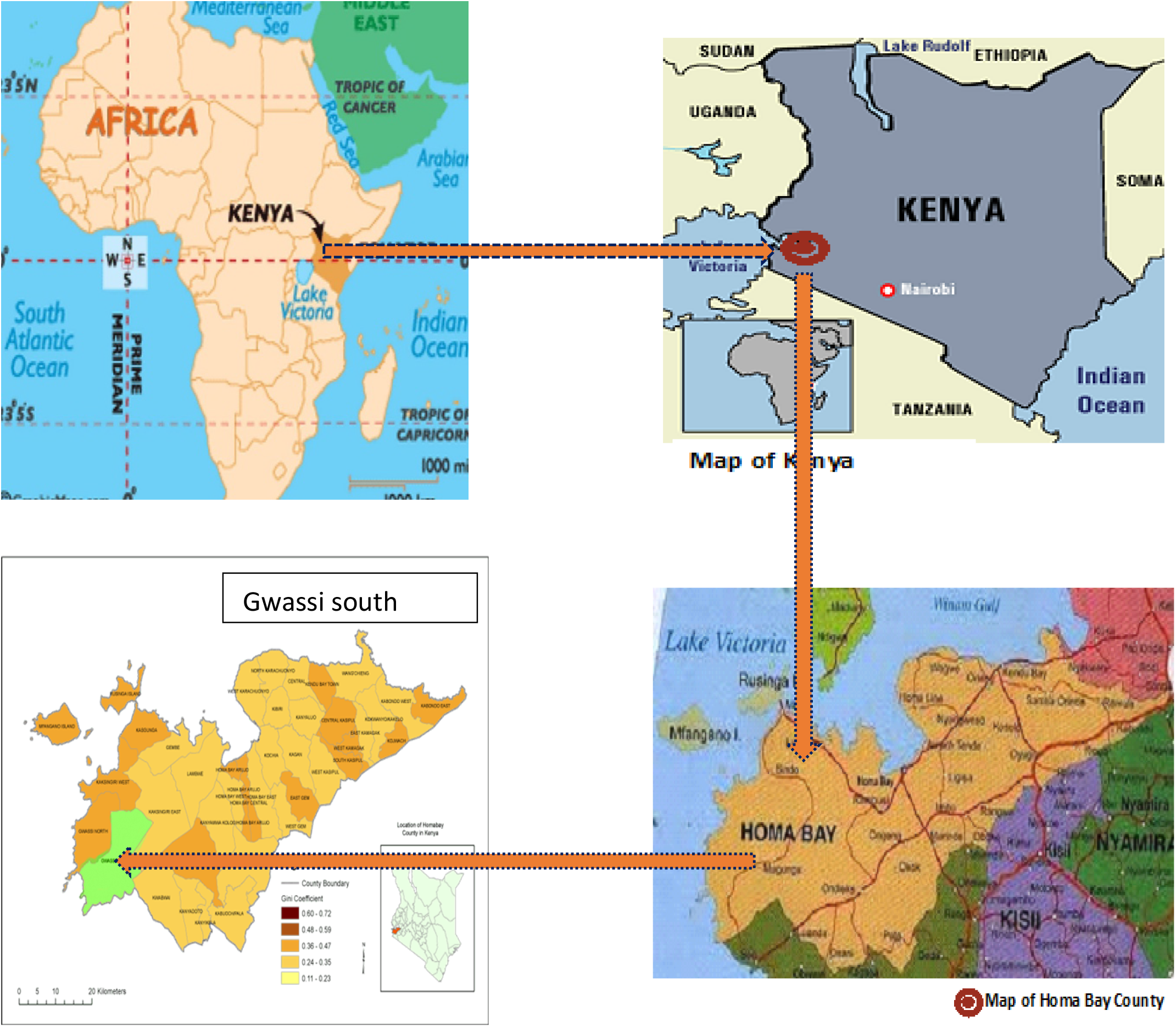
Map showing location of Gwassi South Ward

**Figure 2:**
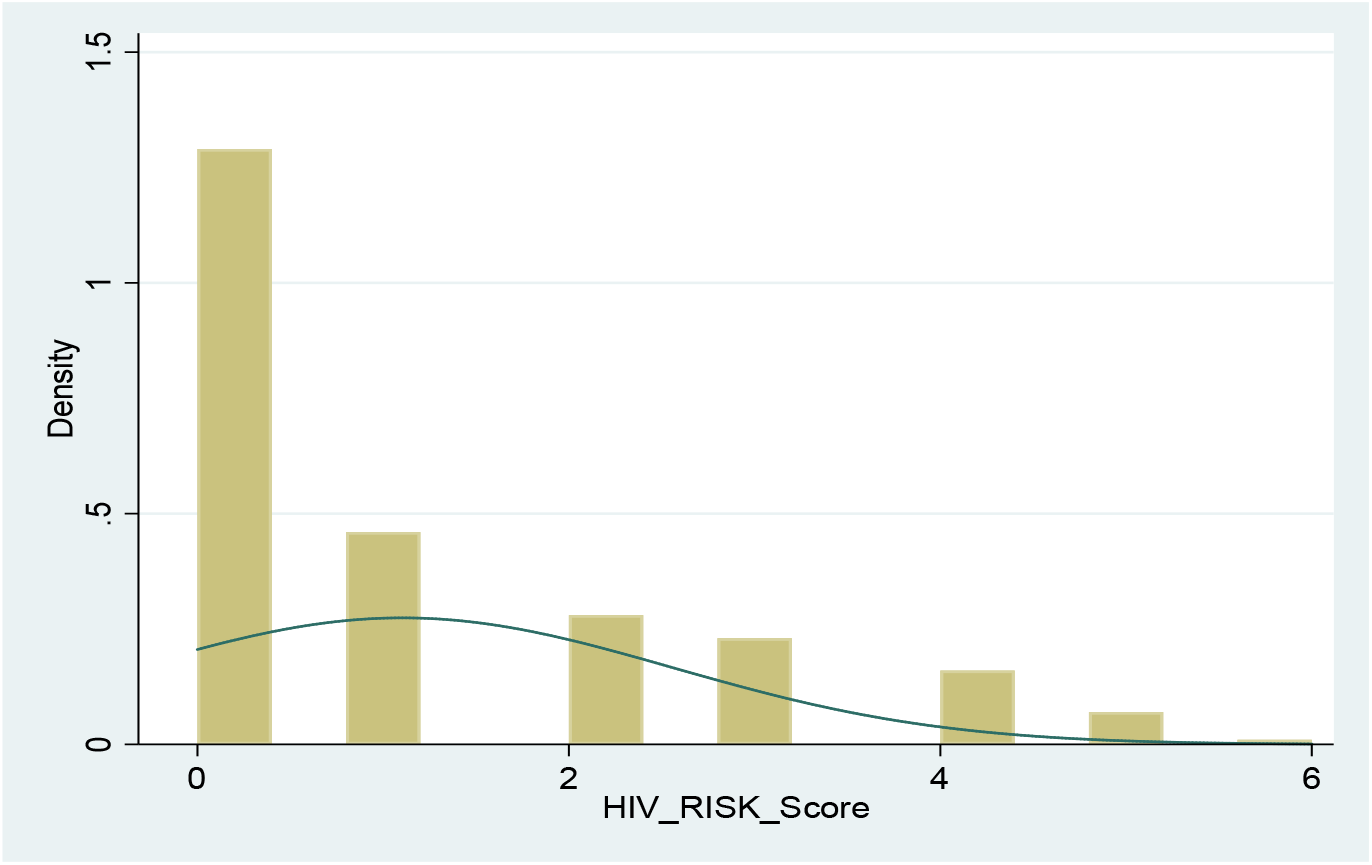
HIV Risk Score of the respondents

### Association of Knowledge about HIV with Sex Practices

The ‘risk’ status, being the dependent variable, was coded as a binary outcome, with “Low Risk” coded as “0” and “High Risk” coded as “1”. The knowledge about HIV in categories of “low”, “moderate” and “high” was the independent variable. The output was shown in the Table 9 below.

**Table 9:**
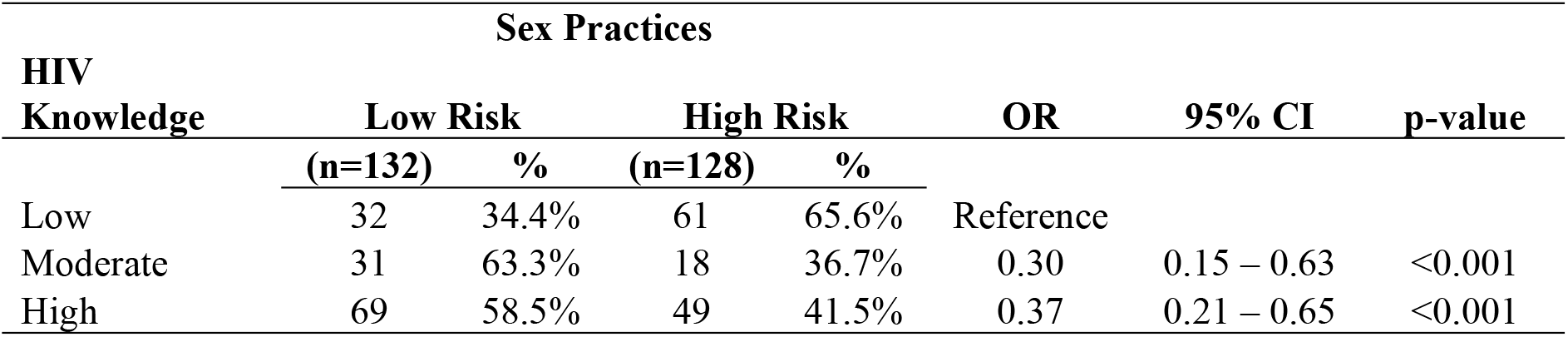
Association of knowledge about HIV with sex practices.

| HIV Knowledge | Sex Practices |  |  |  | OR | 95% CI | p-value |
| --- | --- | --- | --- | --- | --- | --- | --- |
|  | Low Risk |  | High Risk |  |  |  |  |
|  | (n=132) | % | (n=128) | % |  |  |  |
| Low | 32 | 34.4% | 61 | 65.6% | Reference |  |  |
| Moderate | 31 | 63.3% | 18 | 36.7% | 0.30 | 0.15 – 0.63 | <0.001 |
| High | 69 | 58.5% | 49 | 41.5% | 0.37 | 0.21 – 0.65 | <0.001 |

The binary logistic regression analysis (in the table) above shows that respondents with moderate (OR=0.30; 95% CI, 0.15–0.63, p<0.001) and high HIV knowledge (OR=0.37; 95% CI, 0.21– 0.65, p<0.001) were less likely to demonstrate high risk sex practices as compared to respondents with low HIV knowledge.

### Association of attitude about HIV with Sex Practices

In order to establish attitudes on HIV and AIDS towards safe sex practices among students in Secondary Schools, a binary logistic regression was performed. The sex practices status was taken as the dependent variable, coded as a binary outcome, with “safe risk” coded as “0” and “High risk” coded as “1”. The attitudes about HIV in categories of “Positive” and “Negative” was our independent variable (Table 10).

**Table 10:** Association of attitude about HIV with sex practices.

| HIV<br>Attitude | Practices |  |  |  | OR | 95% CI | p-value |
| --- | --- | --- | --- | --- | --- | --- | --- |
|  | Safe |  | Risky |  |  |  |  |
|  | (n=132) | % | (n=128) | % |  |  |  |
| Positive | 101 | 63.5% | 58 | 36.5% | Ref. |  |  |
| Negative | 31 | 30.7% | 70 | 69.3% | 3.93 | 2.31 – 6.69 | <0.0001 |

The binary logistic regression analysis shown (in the table) above revealed that respondents with negative attitude were 4 times more likely (OR = 3.93; 95% CI, 2.31 – 6.69, p<0.0001) to express high risk sex practices as compared to respondents with positive attitude toward HIV and AIDS.

### Strategies on HIV and AIDS

The fourth objective of the study was to establish intervention strategies on HIV and AIDS by secondary schools in Gwassi South Ward in Suba South sub-County. Key informant interviews were conducted with 30 individuals (22 headteachers and 8 health masters) from among the 11 secondary schools in Gwassi South ward. Some of the most striking qualitative data from head teacher interview schedules are shown below. All the 11 schools had the revised HIV curriculum in place. Most (76%) of the key informants had been trained on the curriculum while only 12 (4%) reported having implemented the curriculum in the past one month. The curriculum was found to include sections for HIV knowledge, attitude and practices.

*Some of the teachers in my school are trained on teaching and support of HIV and AIDS in our school. There are about 2-4 teachers who have been trained in HIV and AIDS*. [Interviewee 3]

*In our school we have curriculum quality assurance including HIV and AIDS. In addition, we have teacher appraisal form*. [Interviewee 11]

The qualitative data indicates that the secondary schools in Gwassi South Ward have in place intervention strategies on HIV and AIDS.

## DISCUSSION

This study reports an above average level of KAPs relating to HIV and AIDS. However, misconceptions about the routes of transmission of HIV still remain. In addition, only 61.1% of the students showed positive attitudes towards PLHIV. Almost all survey respondents knew about HIV and AIDS and majority of them could correctly answer questions on HIV transmission and prevention, indicates that the majority had good basic awareness of the issue as seen in other studies (Chory et al., 2021). Respondents also answered majority of questions relating to the main routes of HIV transmission correctly. Similar findings were reported in two studies in Afghanistan and Kazakhstan (Forsyth, 2018). However, there was a lack of understanding about some important points of transmission of HIV, such as the lack of knowledge that HIV can be transmitted through oral or anal sex and beliefs that skin condom works better than latex condom against HIV and that a person can get HIV by sharing a glass of water by someone who has HIV. This indicates that students need correct or accurate information routes of HIV transmission. Again, similar misconceptions have been reported in other studies. The majority of students knew that the use of condoms during sexual intercourse could prevent HIV. Similar findings were reported by previous studies conducted amongst some university students in Afghanistan (Mansoor, Fungladda, Kaewkungwal, & Wongwit, 2008).

Students exhibited mixed reactions to PLHIV. Whilst they displayed positive attitudes on most of the issues, a good proportion expressed discomfort sitting next to being around someone who has HIV in public or private transport. This finding is not unique for Gwassi South Ward Secondary School students. Similar results had been reported in several studies conducted in Ghana, China, Turkey and Iran (Hansson, Stockfelt, Urazalin, Ahlm, & Andersson, 2008; Sallar, 2009). A study in Iran in 2002 among 4641 second-grade high school students reported that about half of the respondents (46%) said that they would not want to sit near PLHIV or to shake hands with them. This may be because students at times express empathy towards PLHIV. However, they still fear that having close contact with them might put them at risk of contracting HIV. For instance, most students felt that it is not safe for a person with HIV to look after somebody else’s children. On the contrary, nearly half of the respondents said they were eager to show compassion towards PLHIV. Discriminating attitudes to adolescent PLHIV might be an obstacle for the efficient propagation of awareness programs (Mansoor et al., 2008), and voluntary counselling and testing for HIV.

Many barriers, such as stigma and discriminatory attitudes (Mansoor et al., 2008), also need to be reduced. Adolescent PLHIV should be equally respected and valued in the society. The Ministry of Education has initiated curriculum instruction models for teaching in secondary schools and to make teachers be examples by providing psychosocial and spiritual care to adolescent PLHIV (Mansoor et al., 2008). Adolescents need targeted counselling about safe practices by avoiding, for example, unprotected sexual relationships and exchange of syringes/needles. Risky sexual practices were also highlighted as another barrier for HIV prevention. Almost one-third of the high school students had a history of sexual intercourse, similar to the findings from other reports (Nigussie et al., 2020; Borneskog et al., 2017).

Although condoms are widely available in pharmacies and public health facilities, and/or are cheap to buy, affordability might still be a factor behind its inconsistent use by students in secondary school. Such behaviours are likely to increase the likelihood of HIV spread.

## Limitations

The finding of this study is subject to a few limitations. First, the findings presented in this study were based on self-reports from participants. Self-reports may suffer some extent of social desirability biases from respondents (Latkin et al., 2017). Although anonymity and honesty were both encouraged during the administration of the questionnaires, it is likely to some extent that the process experienced respondent bias in sensitive questions such as on condom use. Second, the scope of this study was confined to interviewing students enrolled in public secondary schools in Suba South sub-County, Gwassi South ward. Secondary school enrollment was a conglomeration of students originating from other wards or parts of the County, and not necessarily Gwassi South ward. It is possible that localized geospatial variations in perceptions of the students could have affected the responses of 17 students who were found to be coming from outside Gwassi South ward; the extent to which this could have affected our findings is unknown. These two limitations were factored in our analysis plan so as to minimize the margin of error.

## CONCLUSION

In conclusion, misconceptions about HIV routes of transmission as well as prevention and control still exist, despite the students displaying adequate knowledge about HIV and AIDS. Negative attitudes towards HIV and AIDS and risky practices are still common with the youth in secondary schools. Knowledge on HIV and AIDS enhanced safer sex practices. Moreover, negative attitude on HIV and AIDS was jeopardizes safe sex practices among secondary students. Overall, this study demonstrates the link between knowledge, attitude and practice towards HIV and AIDS prevention among secondary school students.

Therefore, there is need to enhance students’ knowledge on HIV and AIDS so as to encourage safer sex practices among them. The students should be supported to have positive attitude towards HIV and AIDS so as to reduce stigma among PLWHIV in their schools. The secondary school administrations should ensure that students understand the risk behavior/practices so as to reduce the rate of HIV infection in the society.

## Data Availability

Data has been submitted

## Data Sharing Statement

The datasets used and/or analyzed during the current study are available from the corresponding author on reasonable request.

## Acknowledgments

We acknowledge all those who accepted to participate in this study, especially the secondary school students of Gwassi South Ward, Homa Bay County, Kenya and school principals for their support during data collection.

## Author Contributions

All authors made a significant contribution to the work reported, whether that is in the conception, study design, execution, acquisition of data, analysis and interpretation, or in all these areas; took part in drafting, revising or critically reviewing the article; gave final approval of the version to be published; have agreed on the journal to which the article has been submitted; and agree to be accountable for all aspects of the work.

